# Impact of Operator Technique Preference on Thrombectomy Reperfusion Outcomes

**DOI:** 10.64898/2026.07.01.26357084

**Authors:** Theja Yelam, Pedro N. Martins, Jaydevsinh N. Dolia, Savio Batista, Jonathan A. Grossberg, Aqueel H. Pabaney, Raul G. Nogueira, Alhamza R. Albayati, Diogo C. Haussen

**Affiliations:** Marcus Stroke and Neuroscience Center, Grady Memorial Hospital / Emory University School of Medicine. Atlanta, Georgia; University of Pittsburgh Medical Center (UPMC) Stroke Institute, University of Pittsburgh School of Medicine, PA

## Abstract

**Background:** Randomized trials have shown comparable reperfusion rates among stent-retriever, contact-aspiration, and combined thrombectomy techniques. We aim to evaluate the association between operator device-selection preference and procedural performance metrics.

**Methods:** Retrospective analysis of prospective data from a comprehensive stroke center. “Preferred technique” was defined as a technique used in >50% of an operator’s thrombectomies. Main exposure: proportion of usage of a given technique by operators in a certain period; primary endpoint: rate of first-pass effect(FPE).

**Results:** 1405 patients fit inclusion criteria. The first time period(2019- mid 2022/n=839) included 4 operators(3 experienced/1 starting practice), with CoT being overall used in 58.9%, SR in 24.4%, and CA in 16.7%. The second(mid 2022-2024/n=566) included 4 total operators(2 experienced/2 starting), with CA reaching 48.2%, CoT 39.8% and SR 12.0%. The distribution of techniques varied between intra-/inter-operators and most(75.0%) had a preferred technique. The technique with the highest FPE rate was never the most used technique. The chances of operators achieving FPE were not dependent on the previous cumulative success for a given technique. Increasing case volume was associated with higher FPE on average, but the learning rate differed by technique and only contact aspiration had a significant learning curve. The parenchymal hemorrhage rates were comparable for individual operators regardless of technique.

**Conclusion:** Neurointerventionists tended to rely on a preferred technique, which did not necessarily lead to superior reperfusion outcomes. The cumulative success with a given technique did not increase the likelihood of attaining FPE in subsequent cases. Among new operators, a learning curve for contact aspiration was observed.

## INTRODUCTION

Mechanical thrombectomy (MT) has revolutionized the treatment of acute ischemic stroke (AIS) due to large vessel occlusion (LVO), significantly improving patient outcomes. Understanding the impact of Neurointerventionists’ experience on procedural outcomes is increasingly important for optimizing stroke care as the number of stroke centers expands globally.

The relationship between neurointerventionalists’ experience and procedural efficiency in MT remains an area of investigation. Studies have demonstrated improved outcomes with increasing center case volumes^1–5^. Additionally, the impact of individual operator experience on MT efficiency appears to be also relevant ^6–8^. Recent guidelines from international organizations have recommended minimum personal experience requirements for performing MT autonomously^9–11^.

Randomized clinical trial data has demonstrated that stent-retriever, contact-aspiration thrombectomy and combined technique approaches are comparable^12–16^. The factors that may influence the preference of operators for a particular technique is multifactorial, including familiarity/experience, technical characteristics, training, institutional factors, clinical data and, anecdotally, from the perception of optimized self-performance. Additionally, the continuously evolving device technology may have an impact on learning curves and procedural performance. This study aims to evaluate the association between operator device-selection preference and MT procedural performance metrics amongst fellowship-trained Neurointerventionalists.

## METHODS

This is a retrospective analysis of prospective data on patients with LVO strokes treated with thrombectomy spanning 2019-2024 in a single comprehensive stroke center. Inclusion criteria comprised consecutive patients with anterior circulation (intracranial internal carotid artery - ICA, middle cerebral artery - MCA M1, M2, or M3 segments, anterior cerebral artery) and posterior circulation occlusions (basilar, and posterior cerebral artery) treated with either contact aspiration, stent-retriever or combination therapy (contact aspiration plus stent-retriever). Tandem occlusions and isolated cervical carotid or vertebral occlusions were excluded. This study was approved by the local Institutional Review Board.

The study duration was dichotomized into two time periods: 2019- mid 2022 and mid 2022-2024. The first period encompassed 4 operators - three experienced (13, 5 and 4 years of past training) and one starting practice, while the second period included 4 operators – two experienced (8 and 7 years) and two starting their practice. Two operators participated in both study periods.

### Definitions and Endpoints

A “preferred technique” was defined when >50% of the thrombectomies performed by an operator during the study period involved a given technique. The main exposure is the proportion of usage of given technique by operators in a certain time period, and the primary endpoint is the rate of FPE, according to the operator’s experience with different thrombectomy techniques. The primary safety endpoint was the impact of technique on parenchymal hematomas type 1 and 2 (ECASSIII criteria^22^) per operator.

## Statistical Analysis

### Descriptive analysis

Descriptive statistics were used to study the baseline characteristics of the cohort. Continuous variables are reported as Median [IQR] and categorical variables are reported as proportions.

### Main analysis

Techniques were ranked by FPE rate and frequency of use per operator. An exact binomial test assessed whether the highest FPE technique was the most commonly used (H_0_: P(preferred) = 0.5). To evaluate the performance for each operator, we used a mixed effects logistic regression with operators as a random intercept and calendar year, cumulative percentage of FPE, number of years of operator experience, thrombectomy technique, IVT, age, and occlusion site as fixed effects.

To evaluate the probability of achieving FPE for a hypothetical new case using a selected technique for a given operator, we used a mixed effects logistic regression with operators as a random intercept and calendar year, number of years of operator experience, thrombectomy technique, IVT, age, and occlusion site as fixed effects. We then refitted this model with the addition of cumulative percentage of FPE and compared models using analysis of variance (ANOVA). A conventional (non mixed effects) logistic regression model with the same fixed effects was also fitted for comparison.

Within each thrombectomy technique, we evaluated whether FPE rates differed between operators using a hierarchical mixed-effects model with operator as a random intercept and technique as both a fixed variable and a random slope, allowing the effect of technique to vary across operators. Model were adjusted for years of operator post-fellowship neurointerventional experience, age, occlusion site, and IVT. Predicted values were visualized using the marginaleffects package. A reduced model including technique only as a fixed variable was pursued for comparison via ANOVA.

### Operator influence on technique-specific FPE

To evaluate whether achieving FPE with a given technique meaningfully depends on which operator performs the procedure, we fitted a hierarchical mixed effects model with FPE as outcome, operator as a random effect, and technique as both a fixed variable and a random slope, allowing both intercepts and technique effects to vary by operator. Pairwise contrasts between techniques and operator specific slopes from this model were used to visualize operator specific differences in technique performance.

For all models, calendar year and number of years of operator experience were modelled to account for temporal trends. Predicted values were visualized using predictions from the Marginaleffects package, displaying the probability of achieving FPE and between operator variability was assessed using intra class coefficient (ICC).

### Secondary analyses

#### Learning curves for new operators

To evaluate whether the rate at which operators improve their FPE (learning curve) differs by thrombectomy technique, we used a mixed effects logistic regression model evaluating FPE as the outcome, and with operator as the random intercept and an interaction between cumulative number of cases and thrombectomy technique as main effect, adjusting for age, occlusion site, IVT, and calendar year. Predicted FPE and marginal slopes (change in FPE rates per additional case) were estimated using the predictions and slopes functions from Marginaleffects package.

The overall number of cases performed with a specific technique per operator was not used as an adjustment variable since it would not take into account the evolution (characterizes an end result); the cumulative percentage of FPE (average rate of FPE with each consecutive procedure) was used instead.

#### Safety analyses

To evaluate parenchymal hematoma rates, a generalized linear model was pursued with an interaction between operators and technique, allowing the evaluation of operator-dependent effects across different techniques, while adjusting for age, IVT, occlusion site, ASPECTS, NIHSS, and number of years of experience with technique using restricted cubic splines. Predicted probabilities of parenchymal hematoma were obtained using the marginaleffects package, and marginal slopes were used to quantify the change in parenchymal hematoma risk with increasing number of cases.

We used R Software v4.5.1 (R Foundation for Statistical Computing, Vienna, Austria) for all analyses.

#### Missing data

Data have missing ASPECTS score for 131 cases and of which 88 are posterior circulation and 43(3.5%) is missing and excluded from safety analysis where ASPECTS is used as adjustment variable. No further variable had significant missingness, therefore we have not used any imputation method.

## RESULTS

### Descriptive Analysis and Technique Preferences

Out of 1612 patients treated within the study period, 1405 fit inclusion criteria. The baseline characteristics of the cohort are depicted on **Supplemental Table 1**.

The first time period (2019 - May 2022, n=839) included 4 operators, with CoT being overall used in 58.9%, SR in 24.4% and CA in 16.7%. The second (May 2022 - 2024, n=566) included 4 total operators, with CA becoming the most commonly utilized technique overall (48.2%), CoT being used in 39.8% and SR declining to 12.0% (**Table 1**). Although the distribution of techniques varied between intra-and inter-operators, most (75.0%) operators had a preferred technique (>50% of usage).

**Table 1:**
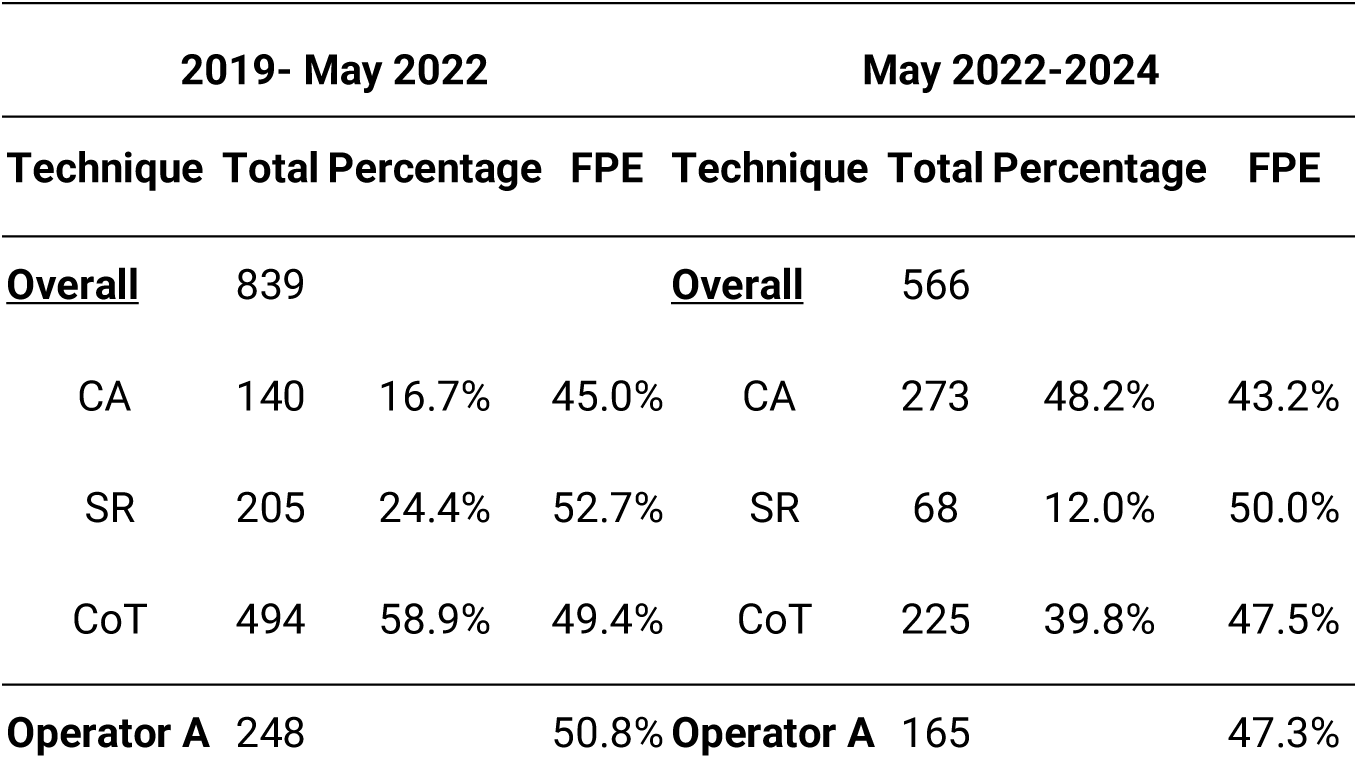
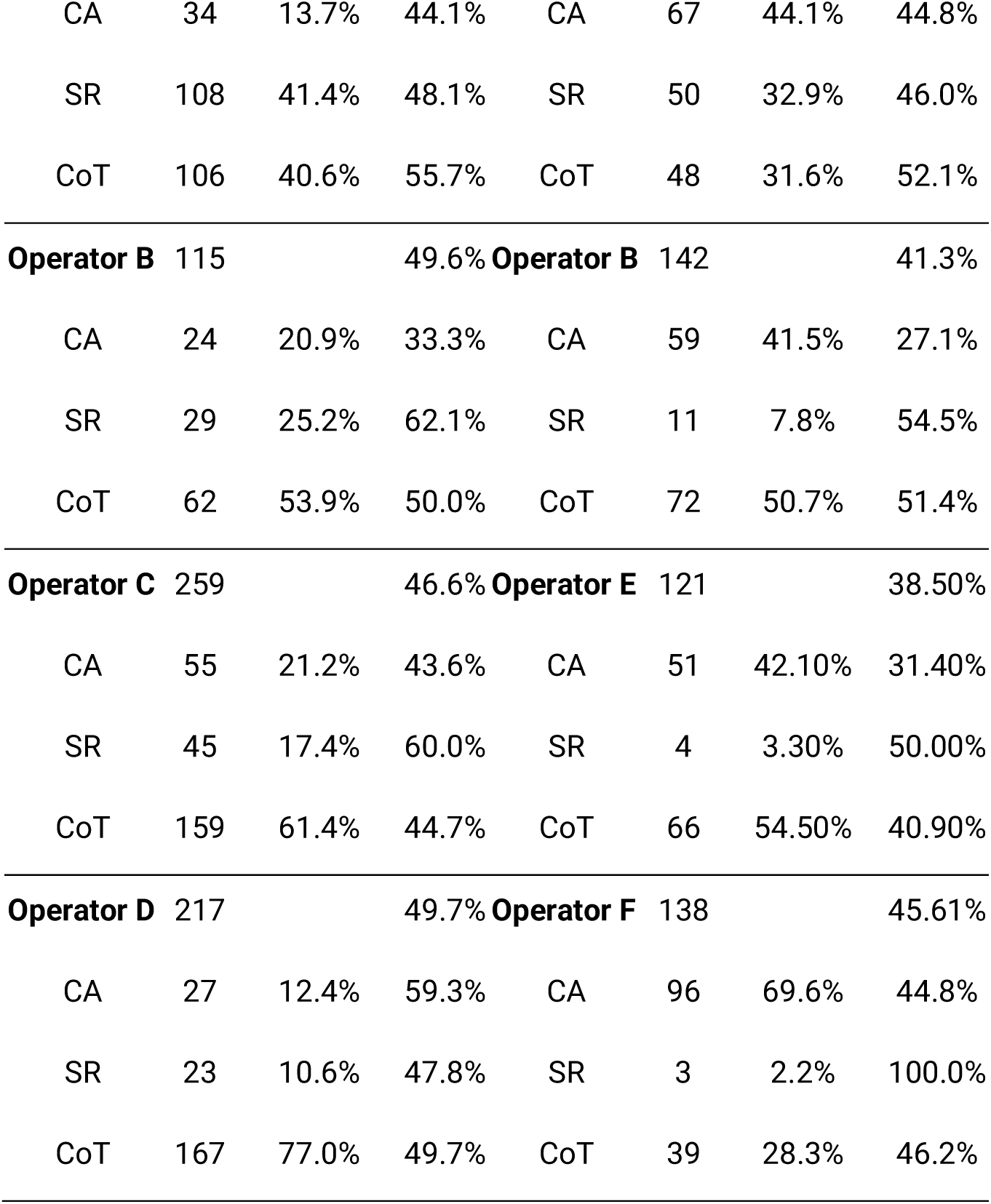
Distribution of operators, number of cases and first-pass effect per technique.

| 2019- May 2022 |  |  |  | May 2022-2024 |  |  |  |
| --- | --- | --- | --- | --- | --- | --- | --- |
| Technique | Total | Percentage | FPE | Technique | Total | Percentage | FPE |
| <u>Overall</u> | 839 |  |  | <u>Overall</u> | 566 |  |  |
| CA | 140 | 16.7% | 45.0% | CA | 273 | 48.2% | 43.2% |
| SR | 205 | 24.4% | 52.7% | SR | 68 | 12.0% | 50.0% |
| CoT | 494 | 58.9% | 49.4% | CoT | 225 | 39.8% | 47.5% |
| **Operator A** | 248 |  | 50.8% | **Operator A** | 165 |  | 47.3% |
| CA | 34 | 13.7% | 44.1% | CA | 67 | 44.1% | 44.8% |
| SR | 108 | 41.4% | 48.1% | SR | 50 | 32.9% | 46.0% |
| CoT | 106 | 40.6% | 55.7% | CoT | 48 | 31.6% | 52.1% |
| **Operator B** | 115 |  | 49.6% | **Operator B** | 142 |  | 41.3% |
| CA | 24 | 20.9% | 33.3% | CA | 59 | 41.5% | 27.1% |
| SR | 29 | 25.2% | 62.1% | SR | 11 | 7.8% | 54.5% |
| CoT | 62 | 53.9% | 50.0% | CoT | 72 | 50.7% | 51.4% |
| **Operator C** | 259 |  | 46.6% | **Operator E** | 121 |  | 38.50% |
| CA | 55 | 21.2% | 43.6% | CA | 51 | 42.10% | 31.40% |
| SR | 45 | 17.4% | 60.0% | SR | 4 | 3.30% | 50.00% |
| CoT | 159 | 61.4% | 44.7% | CoT | 66 | 54.50% | 40.90% |
| **Operator D** | 217 |  | 49.7% | **Operator F** | 138 |  | 45.61% |
| CA | 27 | 12.4% | 59.3% | CA | 96 | 69.6% | 44.8% |
| SR | 23 | 10.6% | 47.8% | SR | 3 | 2.2% | 100.0% |
| CoT | 167 | 77.0% | 49.7% | CoT | 39 | 28.3% | 46.2% |

### Primary Analysis: Preference versus Performance

Overall, the frequency with which a given technique was used was not associated with higher chance of FPE, as the average change in FPE (per percentage point) with increased use was minimal across all three techniques (OR per percentage point of 1.04 (95% CI 0.99-1.10) for CA, 1.01 (95% CI 0.97-1.05) for CoT, and 1.00 (95% CI 0.93-1.09) for SR).

For each individual operator, the technique with the highest FPE rate was never the most commonly used technique (**Table 1**). The chances of an operator achieving FPE were not dependent on the previous cumulative success for a given technique **(Figure 1A and Supplemental Figure 1A)** (adjusted mixed effects model with and without cumulative percentage of FPE showed no significant difference; χ2=6.46, df = 3, p = 0.09, **Supplemental Table 2**). There was limited between operator heterogeneity in terms of FPE rates across different techniques, with an estimated operator variance was 0.04 (SD = 0.21), (**Figure 1B, Supplemental Table 3)**. There was no evidence that the effect of technique on FPE rates is influenced by which operator performs the procedure (LRT for operator specific technique slopes p=0.90; **Figure 1C)**; additionally, there was negligible between-operator performance in comparative relative technique analyses [difference in predicted probability of 0.001 for CoT (CI: 0.000-0.039), 0.017 for SR (CI:0.000-0.211) and 0.023 for CA (CI:0.000-0.130)].

**Figure 1A.**
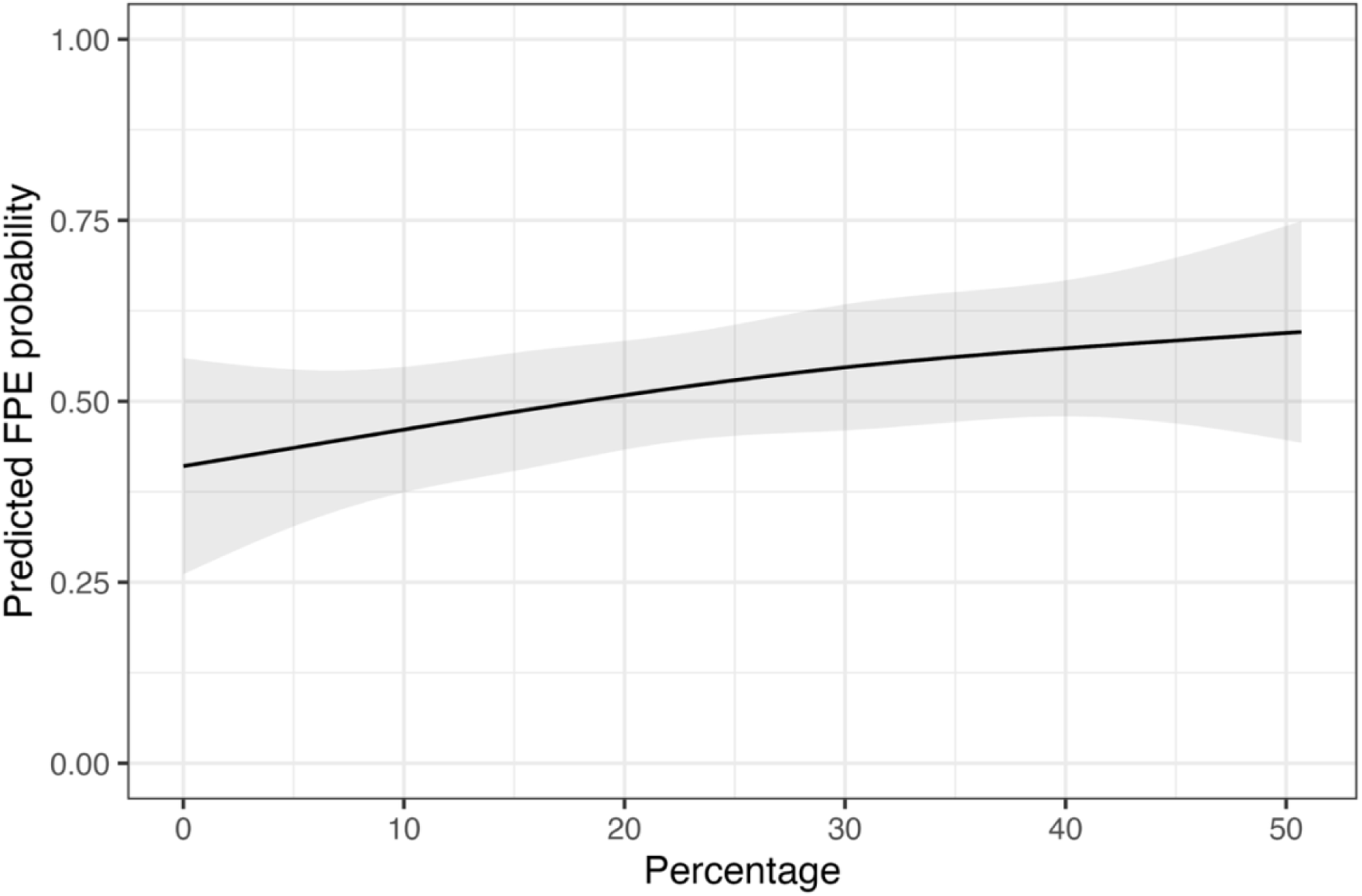
Cumulative percentage of FPE for operators (mixed effects model). FPE, First Pass Effect; Percentage, Cumulative percentage of FPE

**Figure 1B.**
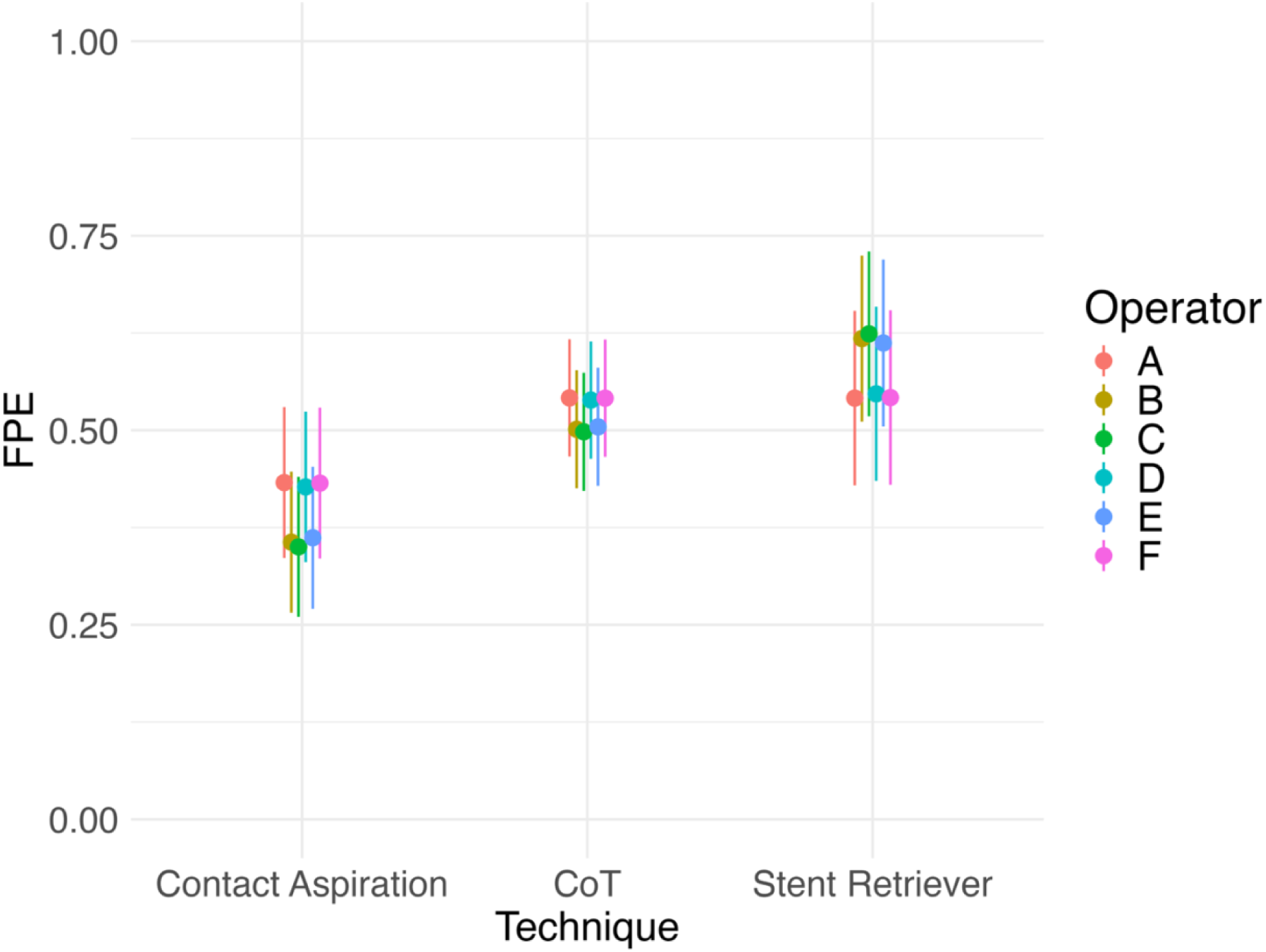
Operator variability within techniques. FPE, First Pass Effect; CoT, Combined Technique

**Figure 1C.**
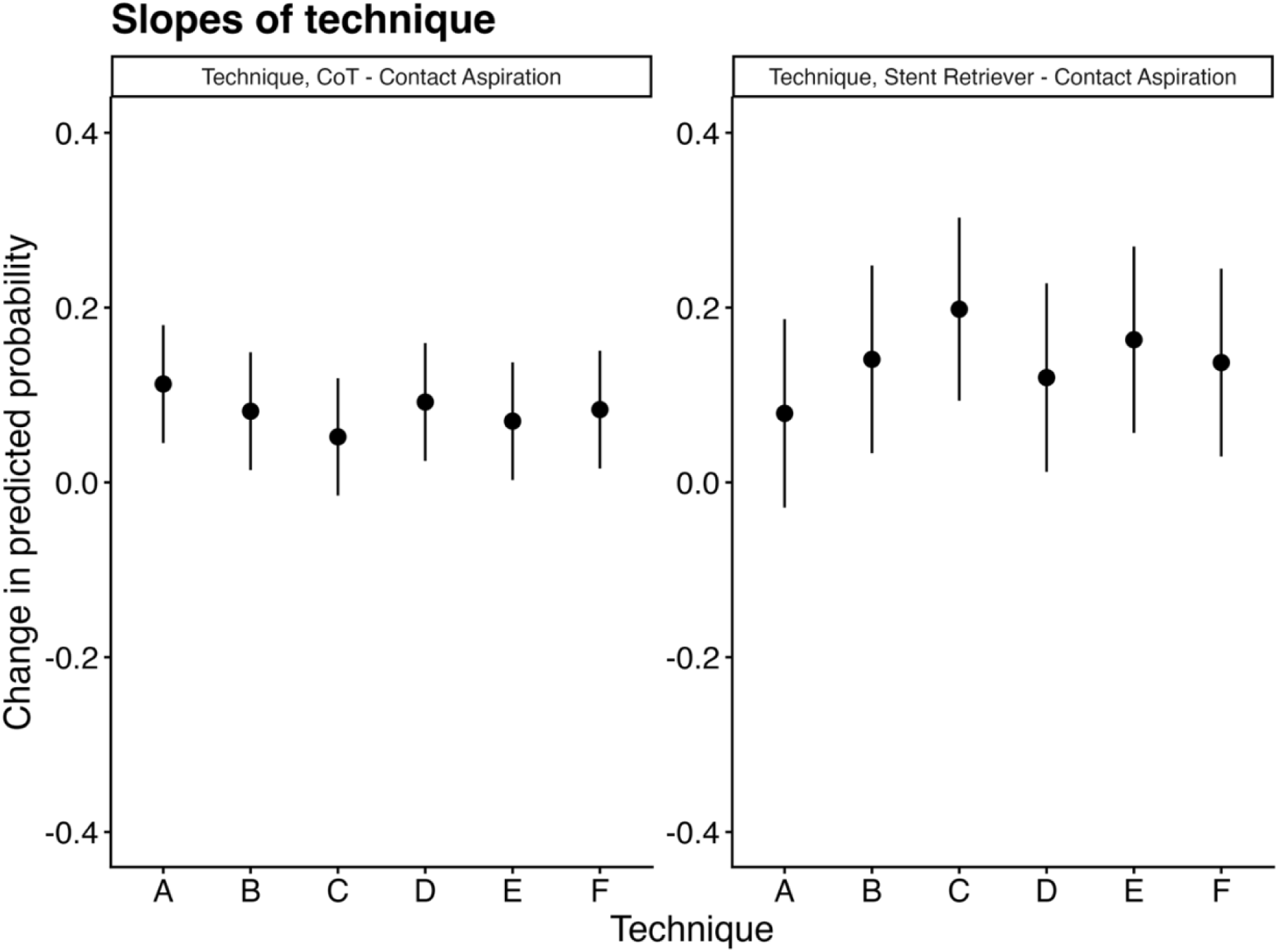
Operator influence on a technique-specific FPE. CoT, Combined Technique

**Figure 1D.**
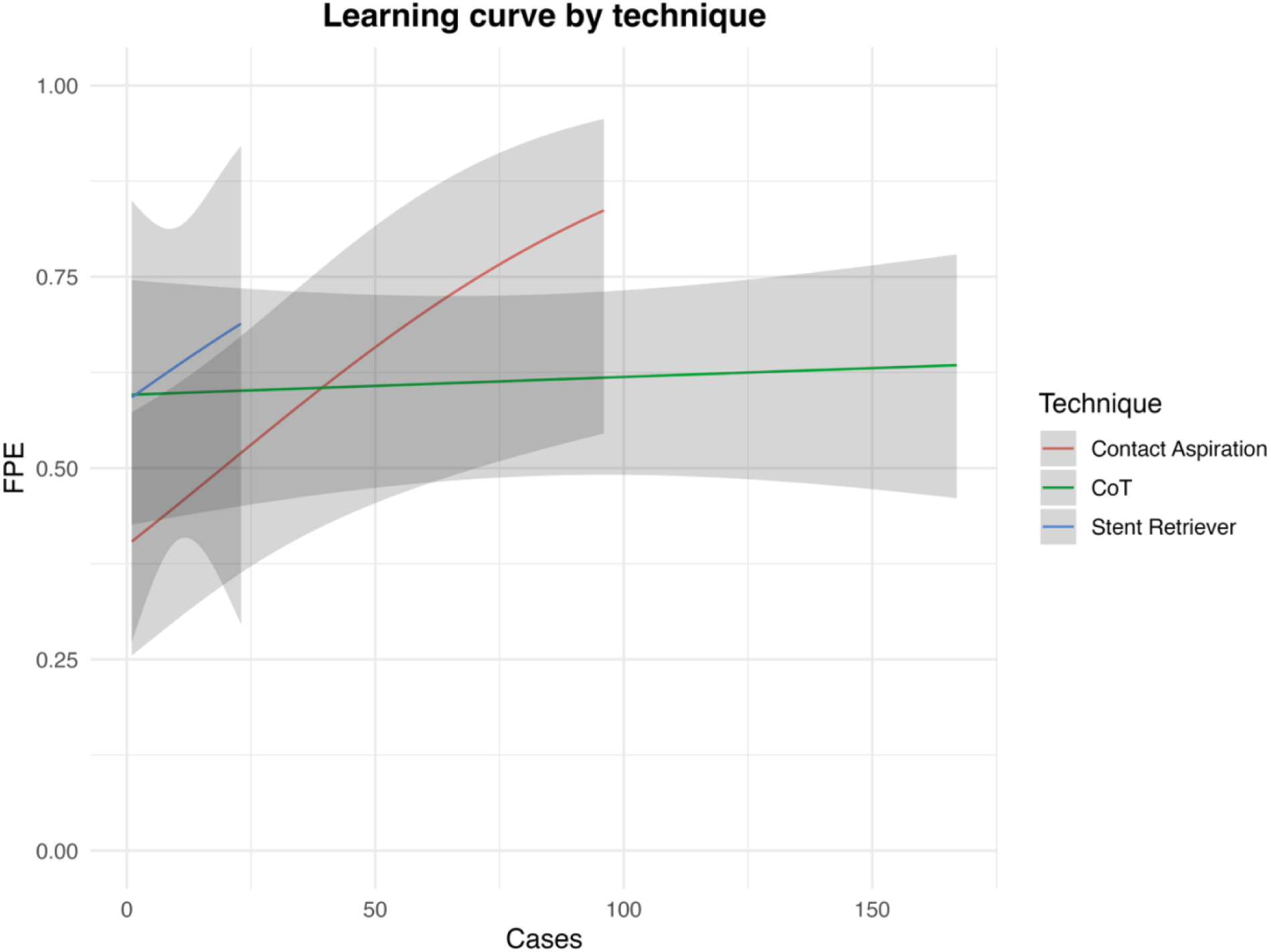
Learning Curve by technique for new operators. FPE, First Pass Effect; CoT, Combined Technique

### Secondary analyses

#### Learning curves for new operators

Within new operators, an overall increase in case volume was associated with higher FPE (β = 0.021, p = 0.006). When examining change in reperfusion performance with increasing case volume for isolated specific techniques, we were only able to demonstrate that contact aspiration had a significant learning curve, with FPE improving as operators gained experience (slope for CA: 0.004, p < 0.01, **Figure 1D**). Whereas SR and CoT showed no significant association between case volume and FPE (slopes for SR: 0.004, p = 0.7, and CoT: 0.0002, p = 0.7; **Figure 1D)**, although the SR sample for new operators was small. The comparative analysis between CA and CoT showed that the per-case improvement in FPE was smaller for CoT (CA vs CoT β = - 0.020, p = 0.02), while was not meaningfully different for CA vs SR (CA vs SR β =-0.002, p = 0.97). There is no detectable variability between new operators in achieving FPE (operator variance is 0; singular fit).

#### Safety analysis

For individual operators, the rates of parenchymal hematoma did not significantly differ across thrombectomy techniques (pairwise operator-level comparisons, all p>0.05).)(**Supplemental Table: 4 and Supplemental Figure: 1B**). The only exception was a lower rate of parenchymal hematoma for SR compared to CA for Operator F, although the sample of SR was very small (n=3).

## DISCUSSION

This single high-volume center involving operators with variable experience and different time periods shows that the chances of FPE were not higher with the preferred technique. Moreover, the cumulative success when using a given technique were not found to influence the chances of achieving FPE in the next case. Among new operators, a learning curve for CA was noted when compared to the CoT.

Randomized clinical trials demonstrated comparable rates of reperfusion between using CA and SR^17^, between SR and CoT^13^, as well as between CA and CoT^12^. In the present study, although operators often demonstrated a preference for a specific technique, the technique they most frequently employed did not correspond to the one associated with their highest reperfusion rates. The available evidence shows that increasing operator volume leads to overall better rates of reperfusion;^6,23^ however, the higher experience with a given technique was not associated with better rates of reperfusion compared to other technical approaches. The present data suggest that the familiar notion in clinical practice (where operators favor the technique they believe performs best *in their hands*, despite limited objective verification) may not hold true.

The impact of cumulative experience with different thrombectomy techniques is not well understood. A study based on the early days of SR use through the SWIFT prime randomized clinical trial roll-in and randomized populations indicated that this modality had a rapid learning curve.^18^ Although stent-retriever technology has been more static compared to catheter technology and that retrievers can be deployed passively, it has been demonstrated that some specific retrievers benefit from active deployment techniques, which can be more technically nuanced, and have not been accounted for in the aforementioned analyses^24,25,31^. The initial experience with direct contact aspiration (“ADAPT” technique) reported that catheters contemporary to the study period were easy to navigate intracranially and led to efficient time metrics^26^. The shorter procedural times using CA may have led to the impression that “*compared with a SR, aspiration is technically easier*”^27^. A relatively small study evaluating the learning curve of different techniques demonstrated that the SR group showed more rapid improvement in procedure times compared to the CA group^28^. Another report from the ETIS Registry demonstrated a positive effect of increasing operator’s experience on the rate of FPE using CA, while this was not observed for SR or CoT. Our study corroborates these findings and indicates that despite the technological improvements in device engineering during the study period, CA appears to have a more substantial learning curve as compared to CoT (the relative low utilization of retrievers by new operators precludes drawing conclusions related to this technology). Considering that the reported FPE rate among super large bore catheter series varied from 42%-72%,^19–21^ it is possible that the advent of superbore catheters may enhance the complexities of CA technique optimization. Additionally, other emerging technologies (such as delivery assist catheters and non-continuous aspiration) could also influence the learning curve^29,30^.

Randomized control trials have shown that safety profiles, including hemorrhagic complications, parenchymal hematoma, and subarachnoid hemorrhage among different techniques are comparable^13,17^. We did not observe safety concerns, particularly not observing lower rates of hemorrhagic complications with more frequently used techniques.

The present study has multiple limitations. It was a single center study without core lab imaging adjudication. As the aspiration catheter technology has substantially evolved over the years as well as the associated delivery techniques, it is hard to appropriately account for its impact on the rates of FPE and the learning curve for different techniques. Even though some operators have very little exposure to a particular technique, and despite having good reperfusion numbers, the use of SR decreased proportionally over time and some of the sub analyses may be underpowered. We did not control for potential differences in stroke etiology, clot composition, or operator technique, which may have influenced technique selection. Considering that all new operators were fellowship-trained, they had a non-quantified exposure to the techniques, which may have influenced the results. It is possible that some operators favor certain techniques based on anatomical features, clot/embolus characteristics, and presumed stroke etiology which would have introduced bias.

In conclusion, operators tended to rely on a preferred technique, which did not confer an advantage in leading to superior reperfusion outcomes. The cumulative success with a given technique did not increase the likelihood of attaining FPE in subsequent cases or to decrease the chances of hemorrhagic complications. Among new operators, a learning curve for contact aspiration was observed.

## SUPPLEMENTAL MATERIAL

### METHODS

#### Statistical Analysis

In all mixed effects models, operators were included as random intercepts to allow the operators to have a different baseline. Calendar year and number of years of operator experience were modelled using restricted cubic splines with 3 knots placed at the 10th, 50th, and 90th percentiles of the distribution and cumulative percentage of FPE was modelled using restricted cubic splines (rcs) with 4 knots at their 5th, 35th, 65th, and 95th percentiles. These specifications were chosen to flexibly capture potential non-linear relationships with FPE without imposing linearity assumptions.

## Data Availability

Will be provided upon resonable request

---

**Supplemental Figure 1A.**
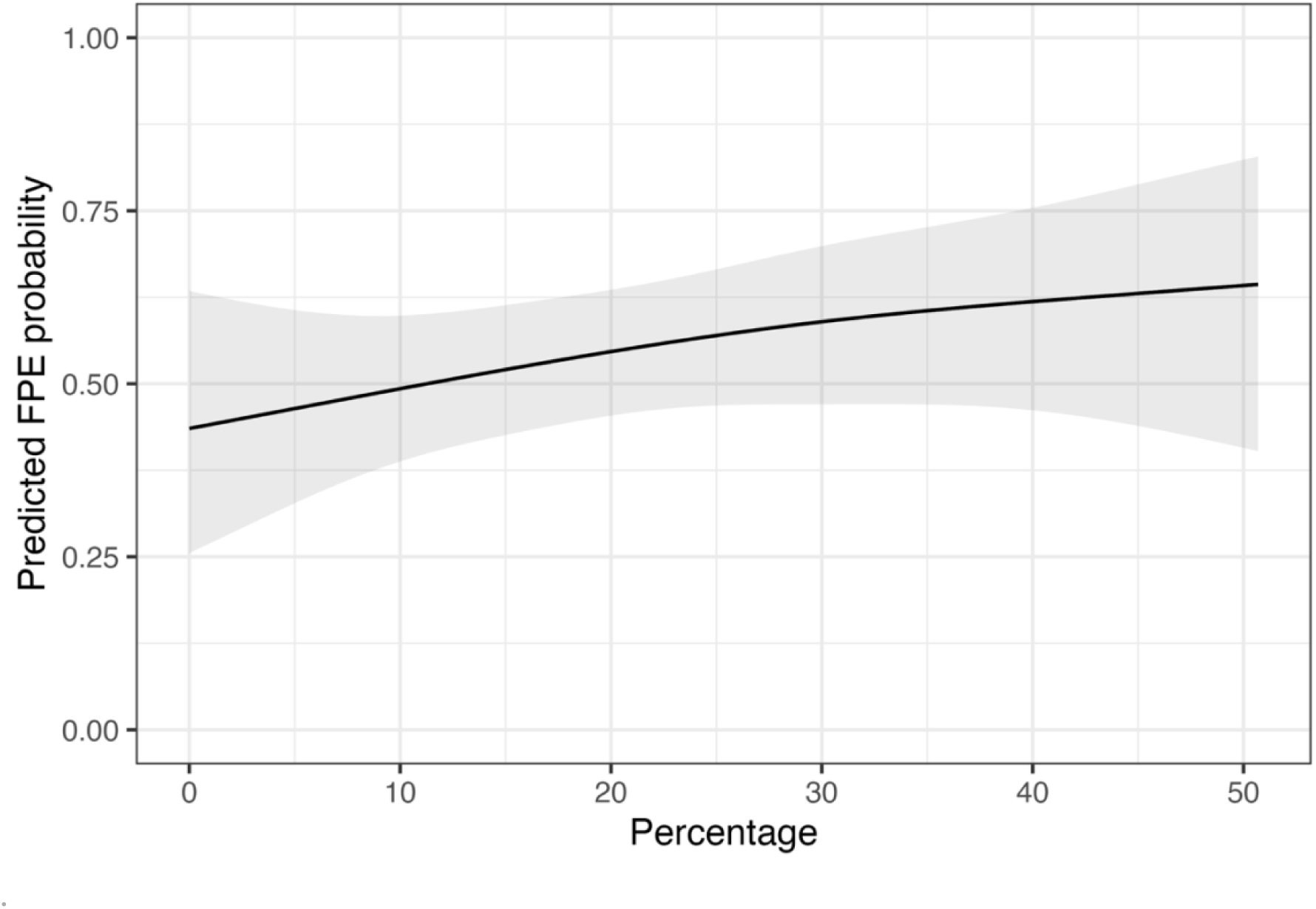
Cumulative percentage of FPE first-pass effect for operators (Linear model); Percentage, Cumulative percentage of FPE.

**Supplemental Figure 1B.**
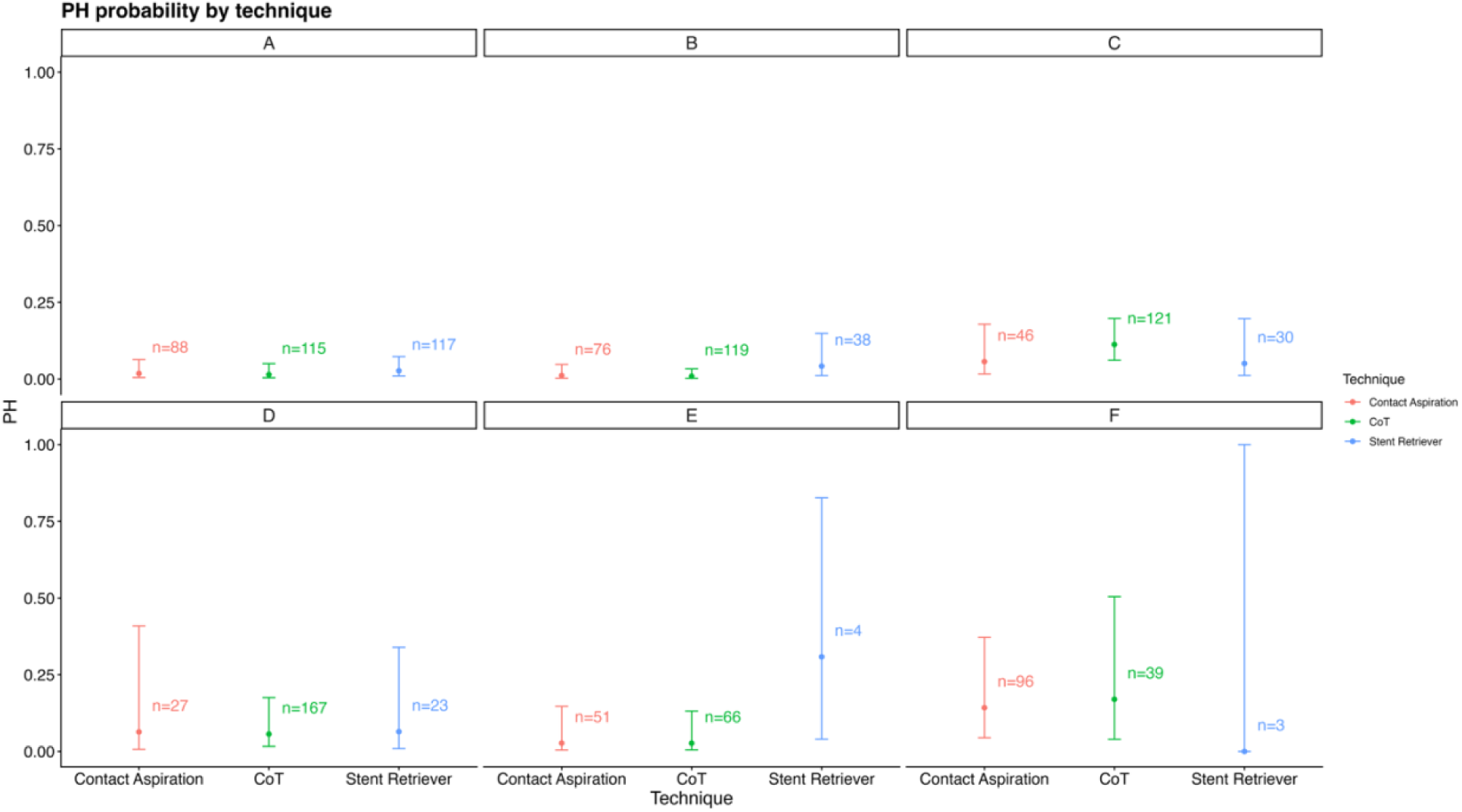
Rates of parenchymal hematoma for each operator across techniques; PH, Parenchymal Hematoma; CoT, Combined Technique

**Supplemental Table 1:** Basic Characteristics.

|  |  |
| --- | --- |
|  | n = 1405 |
| Age [years] | 68 [58-78] |
| Sex |  |
| Male | 744 [53.0%] |
| Female | 661 [47.0%] |
| Ethnicity |  |
| Black | 696 [49.6%] |
| **White** | 470 [33.4%] |
| **Hispanic** | 45 [3.2%] |
| **Asian** | 28 [2.0%] |
| **Other** | 57 [4.0%] |
| **N/A** | 109 [7.8%] |
| **Hypertension** | 1031 [73.3%] |
| **Dyslipidemia** | 344 [24.5%] |
| **Diabetes** | 370 [26.3%] |
| **Atrial Fibrillation** | 402 [28.6%] |
| **Smoking [Current]** | 252 [17.9%] |
| **NIHSS** | 17 [12-22] |
| **ASPECTS** | 8 [7-9] |
| **IV Thrombolysis** | 417 [29.7%] |
| **Anesthesia** |  |
| **GA** | 375 [26.7%] |
| **MAC** | 1030 [73.3%] |
| **Access** |  |
| **Femoral** | 1335 [95.0%] |
| **Radial** | 70 [5.0%] |
| **Occlusion Site** |  |
| **Internal Carotid Artery** | 227 [16.2%] |
| **MCA M1** | 669 [47.6%] |
| **MCA M2** | 351 [25.0%] |
| **MCA M3** | 35 [2.5%] |
| **Basilar Artery** | 88 [6.3%] |
| **Anterior Cerebral Artery** | 19 [1.4%] |
| **Posterior Cerebral Artery** | 15 [1.0%] |
| **mFPE** | 882 [62.8%] |
| **FPE** | 660 [47.0%] |
| **Final TICl 2c-3** | 1063 [75.6%] |
| **Hemorrhage** |  |
| **PH1** | 73 [5.2%] |
| **PH2** | 52 [3.7%] |
| **SAH** | 60 [4.3%] |
| **DC mRS** | 481 [34.2%] |
| **90d mRS0-2** | 462 [32.9%] |
| **Mortality** | 258 [18.3%] |

**Legend:** NIHSS - National Institutes of Health Stroke Scale, ASPECTS – Alberta Stroke Program Early CT Score, MCA – Middle Cerebral Artery, TICl – Thrombolysis in Cerebral Infarction, FPE – First Pass Effect, mFPE – modified First Pass Effect, PH - Parenchymal Hematoma, SAH - Subarchanoid Hemorrhage, mRS – modified Rankin Scale

**Supplemental Table 2:** Model fit statistics and likelihood ratio tests.

| Model | npars | AIC | BIC | logLik | -2*log(L) | Chisq | Df | Pr(>Chisq) |
| --- | --- | --- | --- | --- | --- | --- | --- | --- |
| Model without cumulative percentage of FPE | 21 | 1660.7 | 1767.9 | -809.3 | 1618.7 |  |  |  |
| Model with cumulative percentage of FPE | 24 | 1660.2 | 1782.7 | -806.1 | 1612.2 | 6.46 | 3 | 0.091 |

**Legend:** FPE - First Pass Effect, AIC - Akaike Information Criterion, BIC - Bayesian Information Criterion

**Supplemental Table 3:** Model fit statistics and likelihood ratio tests.

| Model | Random effects structure | Key random-effect variance (log-odds) | SD (log-odds) | AIC | LRT vs simpler model ($\chi^2$, df, p) | Interpretation |
| --- | --- | --- | --- | --- | --- | --- |
| Technique random intercept + slopes by operator | (1 + Technique \| Operator) | Operator intercept: 0.046; CoT slope: 0.011; SR slope: 0.179 | 0.21; 0.11; 0.42 | 1668.3 | $\chi^2 = 2.31$, df = 5, p = 0.80 [from ANOVA] | Added random slopes do not improve fit; estimates unstable (singular fit, Hessian warnings). |
| Technique fixed effect + operator random intercept | (1 \| Operator) | Operator intercept: 0.00000000047 | 0.000021 | 1660.7 | Reference model | Between-operator variance essentially zero; random intercept can be omitted without loss of fit. |

**Supplemental Table 4:** Model Statistics showing parenchymal hematoma rates comparing between techniques for each operator.

| Contrast | Operator | $\Delta\text{Pr(PH)}$ | Std. Error | z | p-value | S | 95% CI |
| --- | --- | --- | --- | --- | --- | --- | --- |
| CoT - Contact Aspiration | A | -0.012 | 0.036 | -0.33 | 0.742 | 0.4 | [-0.083, 0.059] |
| Stent Retriever - Contact Aspiration | A | 0.028 | 0.038 | 0.75 | 0.454 | 1.1 | [-0.046, 0.103] |
| CoT - Contact Aspiration | B | -0.011 | 0.032 | -0.34 | 0.733 | 0.4 | [-0.073, 0.051] |
| Stent Retriever - Contact Aspiration | B | 0.107 | 0.081 | 1.31 | 0.190 | 2.4 | [-0.053, 0.266] |
| CoT - Contact Aspiration | C | 0.077 | 0.060 | 1.28 | 0.200 | 2.3 | [-0.041, 0.195] |
| Stent Retriever - Contact Aspiration | C | -0.009 | 0.072 | - 0.13 | 0.897 | 0.2 | [-0.151, 0.132] |
| CoT - Contact Aspiration | D | -0.009 | 0.078 | - 0.11 | 0.913 | 0.1 | [-0.161, 0.144] |
| Stent Retriever - Contact Aspiration | D | 0.001 | 0.094 | 0.01 | 0.990 | 0.0 | [-0.183, 0.186] |
| CoT - Contact Aspiration | E | -0.001 | 0.042 | - 0.01 | 0.990 | 0.0 | [-0.082, 0.081] |
| Stent Retriever - Contact Aspiration | E | 0.351 | 0.226 | 1.55 | 0.120 | 3.1 | [-0.092, 0.795] |
| CoT - Contact Aspiration | F | 0.030 | 0.077 | 0.39 | 0.696 | 0.5 | [-0.120, 0.180] |
| Stent Retriever - Contact Aspiration | F | -0.173 | 0.041 | - 4.28 | <0.001 | 15.7 | [-0.253, - 0.094] |

**Legend:** CoT - Combined Technique

